# A mixed method study on supportive supervision of Community Health Workers from Central India

**DOI:** 10.1101/2022.01.18.22269339

**Authors:** G Revadi, Ankur Joshi, Abhijit P Pakhare

## Abstract

**Introduction:** Supportive supervision of the Community health workers (CHWs) are critical for their efficient functioning in various health programs. CHWs in India are supervised by facilitators known as ASHA Sahyogini. Our study aimed at investigating the linkage between knowledge and skills of CHW facilitators and their CHWs. To understand the problems encountered by the CHW facilitators while assisting their CHWs. Also, to determine the perceptions of CHWs on nature of supportive supervision of their facilitators.

**Methods:** A mixed method explanatory study using purposive sampling was conducted in a rural block of Madhya Pradesh (Central India). The CHWs were categorised into relatively high performing (RHP) and relatively low performing (RLP) groups based on their performance-based incentives received during (2017-2018). Quantitative component comprised of self-administered questionnaire and skill assessment while the qualitative component comprised of three focussed group discussion with RHP, RLP CHWs and their facilitators.

**Results:** The mean (SD) supportive supervision score given by CHW facilitators were found to be significantly associated with CHWs performance. Problems faced by CHW facilitators while assisting their CHWs resulted from inadequate education of CHWs, insufficient incentives, transport problems and repetitive surveys. While some CHWs perceived to have received good assistance from their facilitators, there were a few who were not dependant on their facilitators and executed duties by self.

**Conclusion:** CHWs performance cannot be ascribed completely to the CHW facilitators performance. The challenges perceived by CHW facilitators were unmodifiable and requires them to strongly motivate and support their CHWs in order to improve their functionality.

## Introduction

Globally Community health worker (CHW) programs are considered critical with respect to achieving health related Sustainable Development Goals (1,2). The concept of CHWs in India was initiated by the National Rural Health Mission (NRHM) to address the health demands of the community. CHWs act as a bridge between community and health system and they enable the reach of primary health care services to the community including educational and mobilization services (3). One CHW is identified in every village in the country, while larger villages have norm of one CHW per 1000 population (4). CHWs receive performance-based incentive (PBI) at the rates decided by state public health administration. At village level, CHWs work in coordination with other cadres of workers like Auxiliary Nurse Midwife (ANM), Anganwadi worker to accomplish the aforesaid tasks.

The CHW program monitoring system has been established to monitor the functionality as well as the health outcomes of the CHWs (5). For this purpose, CHW facilitators were introduced to act as a link between CHWs and the support structure at the block level. One CHW facilitator would supervise 20 CHWs to cover a population of about 20,000. The CHW facilitators serve as essential vehicle for monitoring, supervision and in providing on-site assistance to the CHWs (5). They provide supportive supervision by accompanying their CHWs during village visits, reaching the marginalized people, attending meetings and trainings at different levels, facilitating their selection and addressing their grievance thereby accounting to improve their functionality (5).

Also, CHWs competence are found to be extrinsically influenced by their facilitators as supervision tends to increase their motivation indirectly resulting in better performance (6–9). In order to supervise CHWs, their supervisors must have proficiency in tasks which are performed by CHWs as well as understanding of the context in which CHWs work. However, the role of CHW facilitators in CHW performance has not been explored extensively in the Indian context and this forms the primary basis of our study. Our study aimed at investigating the linkage between knowledge and skills of CHW facilitators and their CHWs. Also, to understand the problems encountered by the CHW facilitators while assisting the CHWs in relation to the perceptions of CHWs on the nature of supportive supervision.

## Methodology

### Study design

An explanatory mixed method study was adopted that included quantitative (self-administered questionnaire) followed by qualitative component (Focussed group discussion) of CHWs and their facilitators.

### Study setting

Raisen district in Madhya Pradesh included seven blocks (as per Census 2011) of which rural Obedullaganj block consisting of 229 villages was chosen considering the operational feasibility.

### Study Population

There were a total of 213 CHWs (as of 2018) that included 189 serving the rural areas and 24 in the urban areas of the block. Data on Performance based incentives (PBI) of the 189 CHWs and the list of their CHW facilitators was retrieved from the Block Programme Management (BPM) unit for the year (April 2017 – March 2018). The population of each of the village served by the 189 CHWs was obtained from the ASHA (CHW database) of which 174 data were available.

### Sample Size and sampling

Purposive sampling techniques was used as there was selective focus on the experiences and perspectives faced by the CHWs with their faciltators and vice versa.

#### a) Quantitative sampling plan

Those 174 villages were classified into three groups based on these population tertiles. Annual PBI of the financial year 2017-18 formed the basis of sample selection. From each group based on the population, all those CHWs who received above 75^th^ percentile of annual PBI were considered as Relatively High Performing (RHP) and those who received below 25^th^ percentile was considered as Relatively Low Performing (RLP).

### Sample Size of CHWs

Out of 174 CHWs in Obedullaganj block, 90 were eligible for participation in the study after adjusting for the population, where only 61 participated in the detailed knowledge and the skill assessment.

#### b) Qualitative component

Focussed Group discussion of the facilitators and the RHP and the RLP CHWs was conducted to identify the determinants of CHWs performance using deductive approach based on preselected determinants from quantitative method and literature review. All their 14 corresponding facilitators were included to study the supervision related factors influencing the performance (High/ Low performance) of the CHWs.

##### Study period

2018 – 2020

## Data collection method and study variables

### 1. Quantitative data collection

CHWs and their facilitators were approached for quantitative data collection at the nearest health facility during their monthly meetings with prior notice through their facilitators at least a week before the data collection. The finalized quantitative questionnaire in the XLS form was entered in ONA software and integrated to the android mobile based ODK 1.19 application. This Self-administered questionnaire in the vernacular language were administered to both the CHWs and their facilitators. The reason being, CHW facilitators were initially working as CHWs for a three-year period subjected to variation in different states of India prior to acquiring the CHW facilitator cadre in addition to their graduate qualification.

#### Study variables

Included questions on socio-demographic details like age, marital status, education, socio economic class, caste, religion, number of family members and number of under 5 children.

#### 1a. Knowledge assessment

There were 25 multiple choice questions which were pooled from different domains of their training modules like maternal and child health, adolescent health and disease control. They were awarded 1 mark for every correct response (no negative markings) and the total knowledge percentage score (out of 25) of every CHW was calculated (Supplementary File 1).

#### 1b. Skill assessment

Skill assessment tool included checklist that contained essential steps to fulfil while performing the home visits during antenatal care with 16 steps, Home based neonatal care with 10 steps and Temperature measurement consisting of 9 steps adopted from the CHW training modules given in (Supplementary File 1). They were awarded 1 mark for every correct step and the total skills percentage score (out of 35) of every CHW was calculated.

#### 1c. Functionality / Supportive supervision score

It is required by CHWs to provide information on the tasks they had performed during that month based on the information at CHW diary to their facilitators with supporting evidence (Supplementary file 2). The reports are then submitted to the officials at block, district, state and then National level, respectively. Also, the functionality score given by the CHW facilitators to their CHWs during every month based on set of tasks (Supplementary File 2) were obtained from the BPM and analysed.

### 2. Qualitative data collection

Three FGDs were conducted separately for CHW fcailitators, RHP CHWs, RLP CHWs on different occasions. The FGD was conducted at the Community Health Centre following prior notice to the concerned authorities a week prior to the meeting. There were a team of 2 interviewers consisting of facilitator for further probing and the other investigator for taking field notes. The probes pertaining to the CHW facilitator and CHWs were different (Supplementary File 1) and those CHWs response pertaining to supportive supervision were enlisted in this paper. The responses were audio recorded using SONY voice recorder.

### Ethics approval

This study protocol was reviewed and approved by Institutional Human Ethics Committee of AIIMS Bhopal (IHEC-LOP/2018/MD0027). Permissions were also obtained from Chief Medical Health Officer, Raisen and Block Medical Officer, Obedullaganj for data retrieval as well as for stakeholder interviews.

Operational definitions are given in - **Supplementary File 3**

### Statistical Analysis

Data in the form of excel sheet was imported from the Ona software and following data cleaning analysis was done using IBM SPSS software version 24 and R software version 4.1.1. Nominal or categorical variables were summarized as frequency and percentage. Continuous variables were summarized as mean and standard deviation when normally distributed and as median and interquartile range when non-normally distributed. For each determinant, association of numerical variable with the binary dependent outcome was done by t-test or Mann-Whitney test as appropriately.

### Qualitative analysis (10)

The data was reviewed through reading of the field notes and active listening of the interview recordings. All the audio-recorded interviews and the handwritten field notes was transcribed. After transcription, data was organized into easily retrievable sections as per topic guides. Each interview in each group was assigned an alphanumerical number. Interviewees were given pseudonyms and other identifiable material was removed from the transcripts. The clean transcript was numbered using line or paragraph numbers. Familiarizing the data was done by means of reading and re-reading making memos and summaries before the commencement of formal analysis. This process was done as major code and minor code manually followed by further categorization. Phrases, sentences or paragraphs from transcript written from voice recorded tapes was utilized as main unit of analysis, and these were arranged physically in groups according to initial coding. Thematic analysis was done that linked both the themes and the sub-themes.

## Results

### Quantitative component

The socio demographic details of the study participants are given in Table 1 where the CHW details are stratified based on the performance (RHP/ RLP). All the CHW facilitators were above 30 years of age and have qualified above primary education with majority 10(71.4%) being graduates and postgraduates. The median age of CHWs were 30(27-35) while that of CHW facilitators were 34(31.5-38).

**Table 1:** Distribution of the Socio-demographic characteristics of CHWs (N=61) and CHW facilitators (N=14)

| Sr<br>N0 | Variables | RLP<br>N=31 | RHP<br>N=30 | Total CHWs<br>N=61 | CHW<br>facilitator<br>N=14 |
| --- | --- | --- | --- | --- | --- |
|  |  | **n (%)** | **n (%)** | **Total** | **n (%)** |
| 1 | Age |  |  |  |  |
|  | Less than or equal to 25 | 9(29) | 0 | 9 | 0 |
|  | More than 25 | 22(71) | 30(100) | 52 | 14(100) |
| 2 | Education |  |  |  |  |
|  | Up to Primary education | 18(58.1) | 7(23.3) | 25 | 0 |
|  | Others | 13(41.9) | 23(76.7) | 36 | 14(100) |
| 3 | Marital status |  |  |  |  |
|  | Married | 29(93.5) | 27(90) | 56 | 14(100) |
|  | Others | 2(6.5) | 3(10) | 5 | 0 |
| 4 | Religion |  |  |  |  |
|  | Hindu | 31(100) | 28(93.3) | 59 | 11(78.6) |
|  | Other religion | 0 | 2(6.7) | 2 | 3(21.4) |
| 5 | Caste |  |  |  |  |
|  | General | 3(9.7) | 7(23.3) | 10 | 5(35.7) |
|  | OBC | 12(38.7) | 11(36.6) | 23 | 8(57.1) |
|  | Scheduled Caste | 4(12.9) | 8(26.6) | 12 | 1(7.1) |
|  | Scheduled Tribe | 12(38.7) | 4(13.3) | 16 | 0 |
| 6 | Socio-economic status |  |  |  |  |
|  | Above poverty line | 15(48.4) | 10(33.3) | 25 | 10(71.4) |
|  | Below poverty line | 16(51.6) | 20(66.7) | 36 | 4(28.6) |
| 7 | No of family members |  |  |  |  |
|  | Less than or equal to 4 family members | 13(41.9) | 10(33.3) | 23 | 10(71.4) |
|  | More than 4 family members | 18(58.1) | 20(66.7) | 38 | 4(28.6) |
| 8 | Under 5 children |  |  |  |  |
|  | Less than or equal to two children | 13(41.9) | 10(33.3) | 23 | 13(92.9) |
|  | More than 2 children | 18(58.1) | 20(66.7) | 38 | 1(7.1) |

Knowledge and skills network diagrams shown in Figure 1 and 2 represents the interconnections between the scores (known as nodes) by CHWs (where RHP CHWs are shown in “green” and RLP CHWs in “orange”) and their facilitators (shown in “blue”) which are related using lines known as edges. Network plot shows interlinkage of every CHW facilitator with their CHWs chosen under the study. The size of each node is directly proportional to the scores obtained by the CHWs and their facilitators.

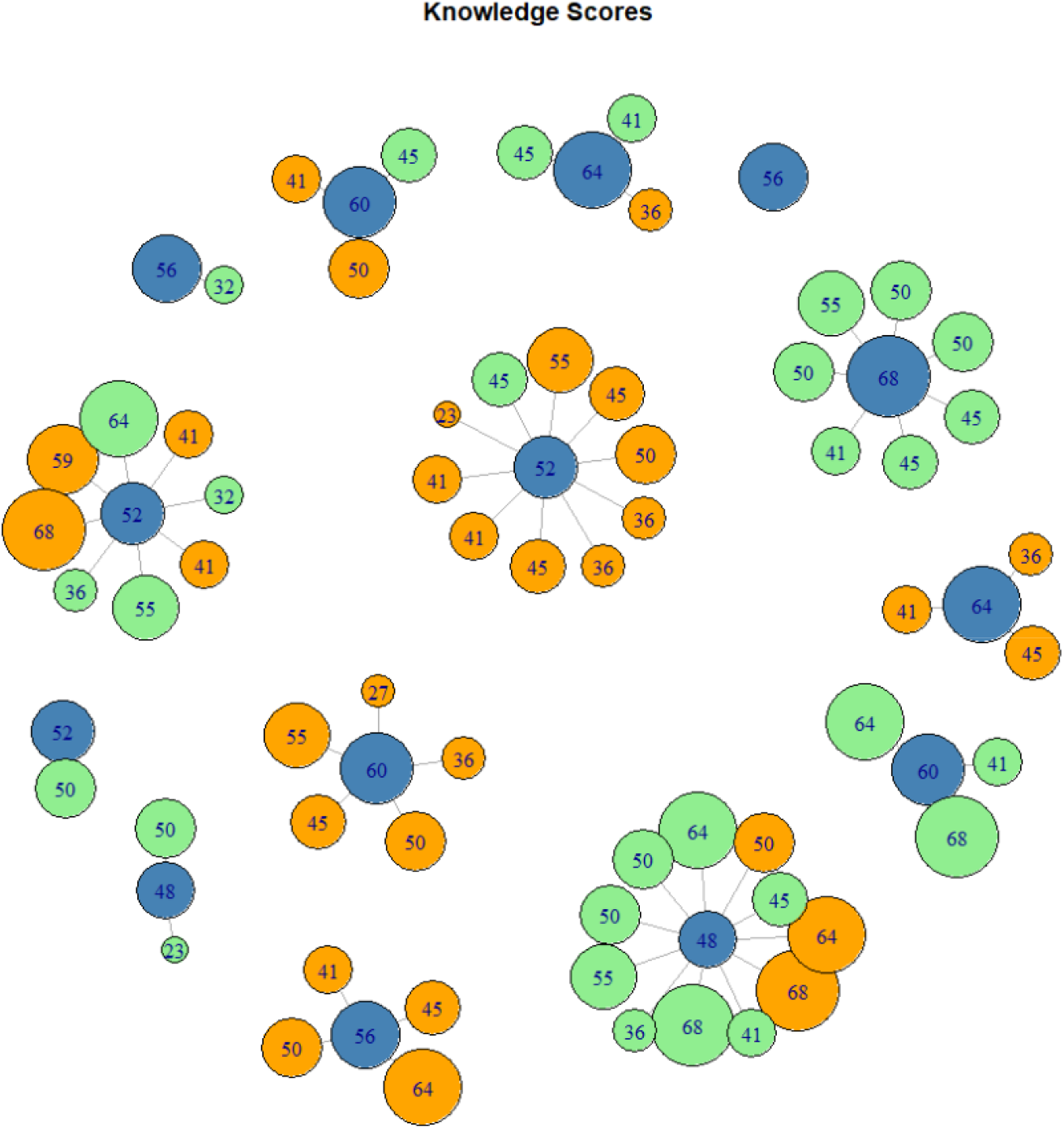

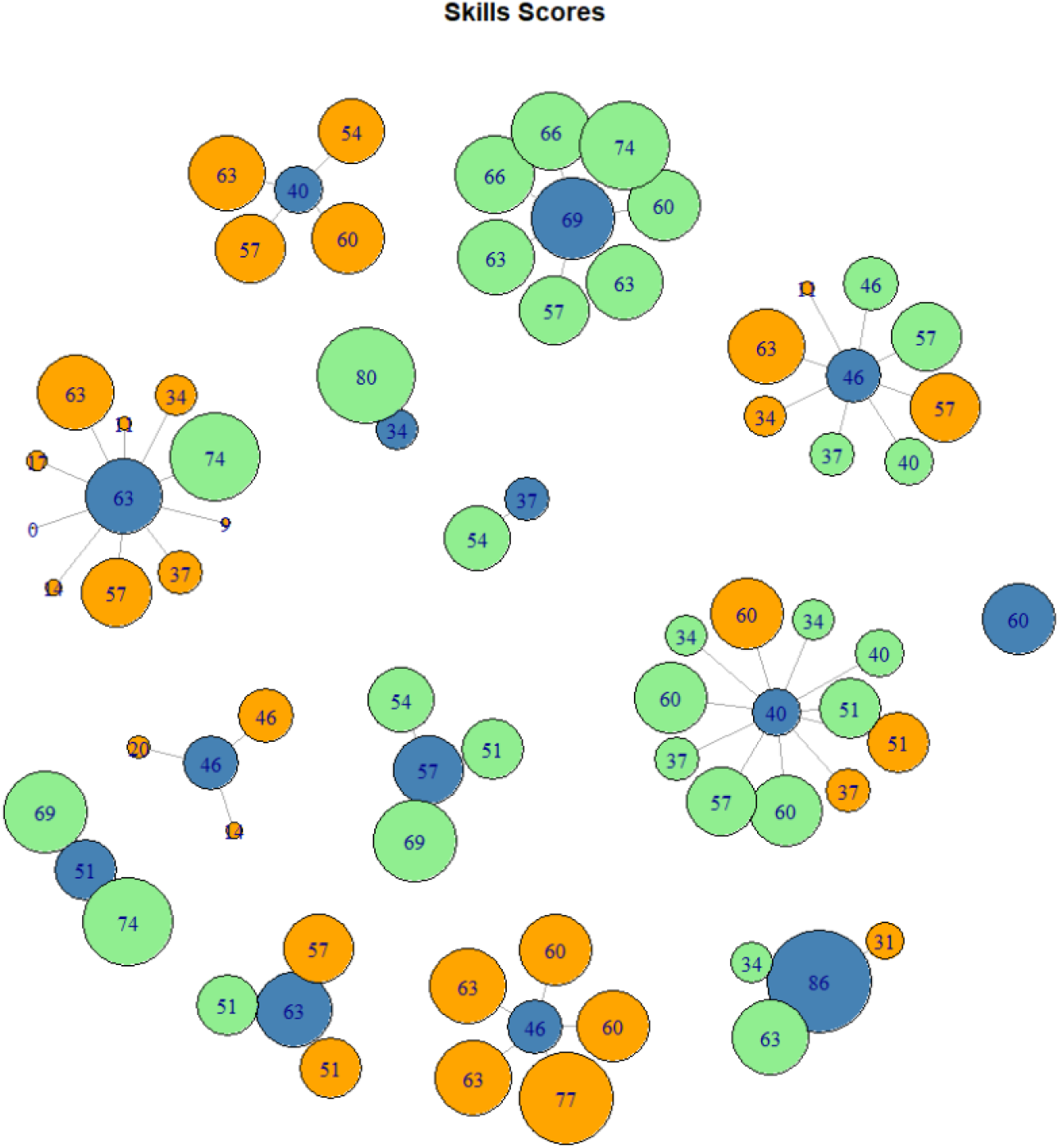

Figure 1 shows that there were only 2(14.28%) CHW facilitators with knowledge score below 50%. Few facilitators were found to have only RHP/RLP CHWs while some had a combination of both. Those CHW facilitators with knowledge score above 50% contained CHWs with above as well as below 50% score. There were instances where some CHW Facilitator had only one selected CHW.

From figure 2, nearly half of the CHW facilitators scored below 50% in their skills. Few facilitators were found to have only RHP/RLP CHWs while some had a combination of both. Those CHW facilitators with knowledge score above 50% contained CHWs with above as well as below 50% score. While those CHW facilitators with below 50% showed relatively higher scores obtained by their CHWs.

The functionality/performance score of CHWs were recorded periodically by the CHW facilitators from 0 to 11 based on the fulfilment of certain definitions of functionality for the each given tasks as given in Supplementary file 2. The mean performance score for the year 2017-2018 was calculated by dividing the total performance score of each CHW by 12 for the given year. The mean (SD) performance score of RLP CHWs were 8.6 (1.42) and RHP CHWs were 9.1 (0.59). The overall mean scores ranged from 0 to 10.5 as displayed in Figure 3. However, the mean performance score given by the CHW facilitator was found to be significantly associated with the performance of CHWs by Mann Whitney test with P value 0.01.

**Figure 3.**
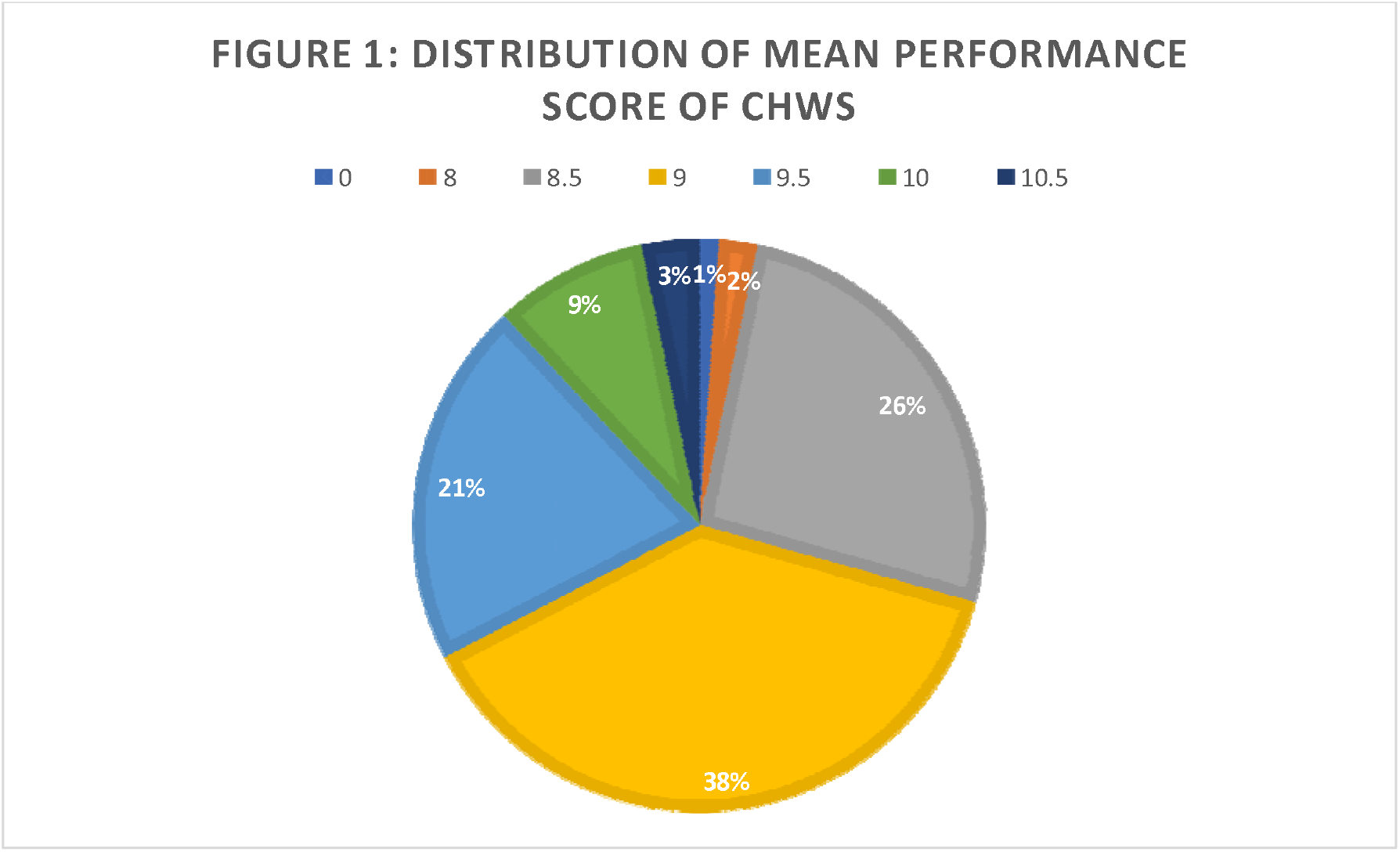
Distribution of the Mean Performance score of CHWs.

### Qualitative results

There were a total of seven CHW facilitators who had participated in the FGD. The perceptions of CHW facilitators on the barriers experienced while supervising and assisting their CHWs were grouped under the themes like Contextual factors, Health System and Individual factors as given below in Table 2.

**Table 2:** Thematic analysis of the barriers experienced by the CHW facilitators while assisting their CHWs:

| Themes | Subthemes | Comments as verbatims |
| --- | --- | --- |
| Individual determinants | Inadequate education | *Those who are well educated do work nicely but still they don't get paid proportionate to their work (Age between 31-35 yrs, PHC).*<br>*While few CHW are less educated, there is a delay in understanding things (Age between 31-35 yrs, PHC).* |
|  | Dependant on family | *When CHWs accompany pregnant women to the hospital, there is no security for her especially at night as she must wait alone for 10-12 hrs. This creates concern among family members as there are children below five years and hence their husband or someone had to bring her all the way from hospital to the village (Age between 31-35 yrs, PHC).* |
| Contextual determinants | Performance is population dependant | *Not every CHW is able to work with equal capacity. Their work is based on the population covered by them. Those who have more population, will get more work to do and get paid more according to their work (Age between 31-35 yrs, PHC).* |
|  | Transport problems | *We are residing in the interior village hence when we call the Janani vehicle, they would say as we are in city now, it would take one hour to reach over there. Hence, delivery tends to happen on the way to the hospital or at home due to delay in the vehicle (Age between 35-40 yrs, PHC).* |
|  | Remote village | *Few CHWs are from interior village and hence they had to come to the main road by walk and then take a vehicle to attend meetings. Hence, they are unable to attend the whole meeting (Age between 35-40 yrs, PHC).* |
|  | Lack of community support | *People residing in the village at times tend to say like : "we will not give you Aadhar id or anything for that matters, why do you ask for it again and again ?" due to repeated surveys by the CHWs (Age between 31-35 yrs, PHC).* |
| Health system | Repeated community | *All surveys don't happen at same time, for example if we consider NCD survey the concerned CHW designated for this* |
| determinants | level surveys | *survey would reply to them as – We have done it already and don't want to do it again (Age between 35-40 yrs, CHC).* |
|  | Less incentives | *CHWs gets paid less than the labourer, so how it is possible to work with such a payment (Age between 35-40 yrs, PHC).* |
|  | Need of Onsite field assistance | *Sometimes we must insist those with difficulty in understanding once / twice or even show them practically then only they can understand those facts (Age between 35-40 yrs, PHC).*<br>*Also, some CHWs do not know to write in their vernacular language so their survey is not reliable at times, so we had to go to the field and solve them (Age between 35-40 yrs, PHC).* |

Two FGDs, one with the 11 RHP CHWs and the other with the 8 RLP CHWs revealed the following feedback on the nature of supervision received from their CHW facilitators. This was questioned to determine the extent of concordance in sharing the drafted responsibilities between the two.

### The following verbatims were by the PHC CHWs based on experiences of supervision from their facilitators

#### Comments on overall supervision by CHWs from PHC area

*“There should be more support from the facilitators. We have no problem in obeying the order of our facilitators, as it is our job to obey the orders. But we receive only less assistance”* **(RHP CHW, age between 31-35 yrs)**.

While other CHW replied *as “ We are not dependant on our facilitators as we do it by ourselves anyways and they are only for the name sake and not for any work”* **(RHP CHW, age between 35-40 years)**.

*One of them commented as “their CHW facilitator helps them, but our facilitator has hardly shown herself up in the last 2/3 months”* **(RLP CHW, age between 41-45 years)**.

#### Experiences on nature of supervision

*“ She only comes during vaccination session or if there is any difficulty in conducting the survey then she comes. There are 14 CHW facilitator out of which only one/ two help us*.*”* **(RHP CHW, age between 35-40 years)**.

Also, *“they call us a night before to inform regarding the submission of forms tomorrow. How is it possible to do it within a short time of notice?”* **(RHP CHW, age between 31-35 years)**.

*“If we have to go for the meeting or do surveys, our CHW facilitator calls us and informs regarding the same prior”* **(RHP CHW, age between 26-30 years)**.

### The following comments were by the CHC CHWs based on experiences of supervision from their facilitators

*“ Our facilitator is very good and helps in almost everything. If I go for mobilising the patients, she even takes care of writing things. She give good support in maintaining our registers* **(RHP CHW, age between 26-30 years***)”*

*“By chance if we don’t go for the meetings in time then she herself comes to our village and collects the forms from us”* **(RHP CHW, age between 31-35 years)**.

For instance, **“***One home delivery had happened in my village after repeated calling of Janani vehicle at least 4-5 times. Our supervisor scolded me for having let the delivery happen at home. Even we were not paid so much that we can arrange a vehicle and bring her to hospital”* **(RLP CHW, age between 26-30 years)**.

## Discussion

The majority of CHW facilitators had better knowledge as compared to their skills. This could be because of their education and years of experience as they were initially CHWs and later became CHW facilitators. Also, other factors like number of trainings and presence of nuclear family with older children could be the potential reasons. Their poor performance in skills could be accounted to lack of availability of the required equipments or forms which leads to inadequate practical exposure over a period. The performance of CHWs cannot be ascribed to their facilitators as it is dependent on other factors like population, education, availability of logistics, number of trainings and nature of supportive supervision. However, the studies on CHW facilitators and their effect on CHWs performance were not explored adequately in literature.

It was also found that functionality score given by the CHW facilitators tends to significantly affect their performance. Similar findings have been observed in other studies (7–9) where supervision was found to enhance CHWs motivation thereby resulting in better performance. Incentives were intrinsic motivators of CHWs performance as evidenced in the studies (11,12). The above findings were the result of population dependant tasks covering various domains like maternal, child health, communicable and non-communicable diseases which form the basis of incentives received by the CHWs. It demands timely submission of the aforesaid task completion number to their facilitator to receive payments.

In our qualitative part of the study, it was found that distance and population tend to significantly affect the performance of CHWs, and this finding was reported in studies (13– 17) where long distances and difficult terrain were found to be associated with poor performance. As the distance increases the frequency of visits to the health care facility tends to decrease and ultimately health seeking behaviour of the community decreases. This scenario tends to worsen when there is unavailability of transport. Hence CHWs of distant villages tends to have comparatively poor performance. Other findings include lower performance among less educated CHWs and those with children below 5 years which was similar to studies (18,19). The possible explanation for these findings could be as there were children below 5 years, it increases the responsibility of CHWs resulting in relatively less time spent on the community due to lactation and other physiological reasons.

There was a mixed nature of supportive supervision where few CHWs provided assistance to such an extent where they had motivated their CHWs and aided them in completion of their tasks efficiently as documented in (5–7) studies. While few CHWs did not receive adequate assistance which could be due to remote village, overburdening of the CHW facilitators as such, lack of interest by CHWs as well as their facilitators.

### Strengths

As it was a mixed method study it was easier to explore all the possible aspects related to supportive supervision from the CHWs and their facilitators in a realistic manner. Not much of studies had been reported focusing on the CHW facilitators thus making it critical to explore. Self-administered questionnaire using mobile based application was employed to prevent interviewer bias which added to the strength of this study.

### Limitations

There were operational constraints for recruitment of larger study participants from different areas therefore this study was conducted at a block level. However, due to limited sample size and the purposive sampling method the generalisability of results cannot be done.

### Scope of further Research

More research should be focussed on the supervision of CHWs as their performance tends to affect the performance of CHWs with respect to frequency of supervision, the quality of supervision amount of time spent, and the contents discussed.

## Conclusion

There were a total of 14 CHW facilitators and 61 CHWs who were covered using mixed method study. CHWs performance cannot be ascribed completely to the CHW facilitators performance however, their supportive supervision score tends to affect their performance. The barriers perceived by CHW facilitators on their CHWs performance were unmodifiable and requires them to strongly motivate their CHWs and aid those in remote areas to achieve effective supervision.

## Data Availability

All data produced in the present study are available upon reasonable request to the authors

## Abbreviations

ASHA: Accredited Social Health Activists
CHWs: Community Health workers
IQR: Interquartile range
NHM: National Health Mission
ODK: Open Data Kit
ONA: Organizational network analysis
RHP: Relatively High Performing
RLP: Relatively Low Performing
XLS: excel Spreadsheet.

## Acknowledgement

Permission and facilitation for data collection at field sites were provided by Block Medical Officer, Obedullaganj block, Raisen District, Madhya Pradesh.

Our sincere thanks to Dr. Deepti Dabar for her valuable guidance and feedback during the process of protocol writing and submission.

We thank our Head of the department, Prof (Dr.) Arun Kokane for his support that had helped us to mobilize the CHWs during our study at various settings.

We thank all the Senior residents, Junior Residents and the interns who helped us during the data collection period and for their cooperation during the study.

## Supporting information

Supplementary File 1: Knowledge and Skill assessment checklist with probes.

Supplementary File 2: Supportive supervision functionality assessment tool

Supplementary File 3 : Operational definitions

## Notes

### Competing Interest Statement

The authors have declared no competing interest.

### Funding Statement

Indian Council of Medical research, New Delhi (ICMR) funded this study under MD thesis grant. The funders had no role in study design, data collection and analysis, decision to publish, or preparation of the manuscript [No.3/2/March-2019/PG-Thesis-HRD (11)].

