## Supplementary figures and images for "A mixed method study on supportive supervision of Community Health Workers from Central India"

### Supplementary File 2

**Supplementary Table 4: Supportive supervision functionality assessment tool** (5)

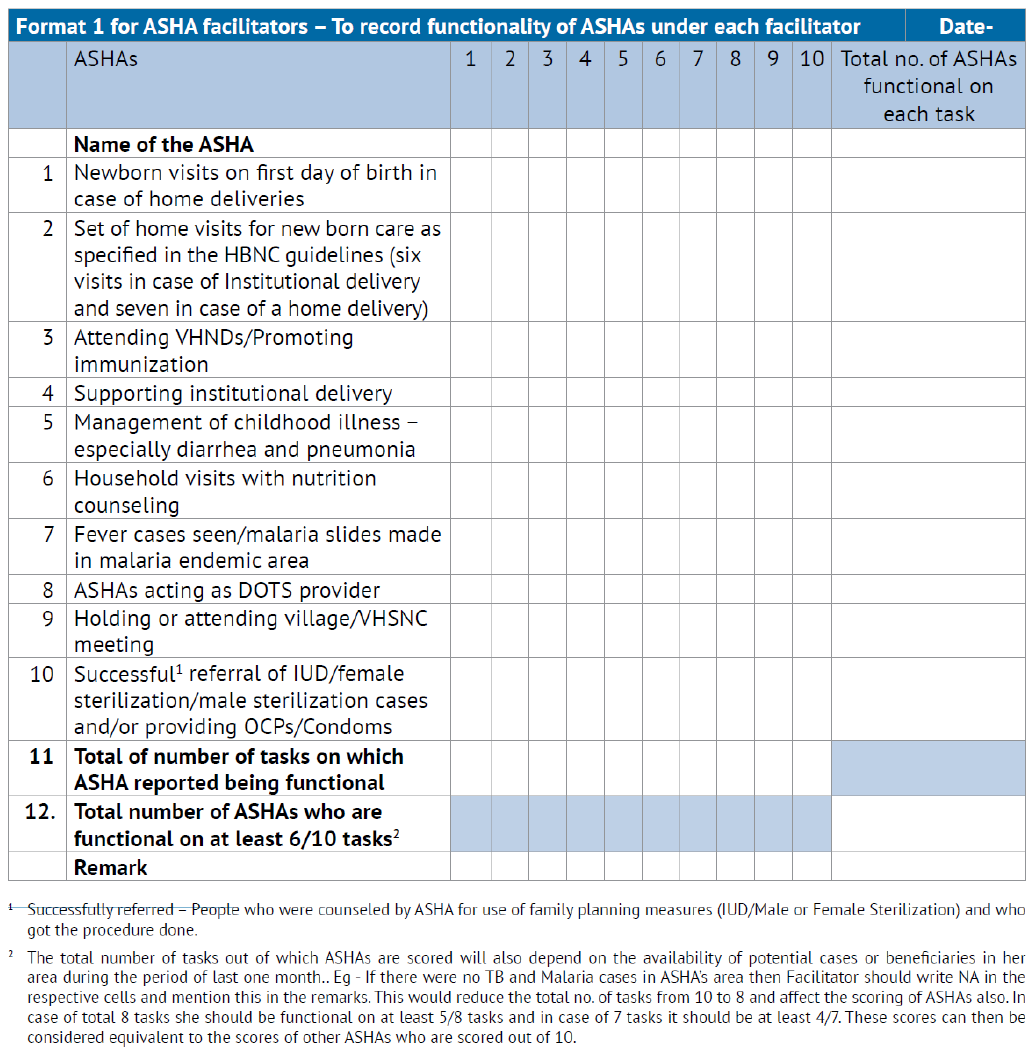
