## Supplementary File 3 for "A mixed method study on supportive supervision of Community Health Workers from Central India"

**Operational Definitions:**

1. ***ASHA:*** CHWs are also popularly known as Accredited Social Health Activists in India.
2. ***National Health Mission:*** The NHM was launched by government of India in 2013 subsuming the National rural health and National Urban health mission. It envisages achievement of universal access to equitable, affordable and quality health care services that are accountable and responsible to people’s need (20).
3. ***Home based newborn care:*** CHWs are mandated to visit every newborn in her area with at least seven visits (Day 1, 3, 7, 14, 21, 28, 42) in case of home-based deliveries and six visits in case of Institutional deliveries (Day 3, 7, 14, 21, 28, 42) (21).
4. ***Urban area*** (22)***:***

(a)  all places with a Municipality, Corporation or Cantonment or Notified Town Area (b)  all other places which satisfied the following criteria:

I. minimum population of 5000
II. At least 75% of the male working population was non-agriculture
III. A density of population of at least 400 sq. Km. (i.e, 1000 per sq. Mile).
5. ***Rural area:*** Those areas that don’t follow the above definition will be included under rural India which accounts to 68.7% (22).
6. ***Village:*** In the rural areas the smallest area of habitation, viz., the village has a definite surveyed boundary, and each village is a separate administrative unit with separate village accounts. It may have one or more hamlets. The entire revenue village is one unit (22).
7. ***Block:* :** India is a large country comprising of 28 states and 8 union territories as per 2011 Census. These states and the union territories are divided into districts. Each district has 6 sub-divisions one of which is a block comprising of 80,000 to 1,00,000 population or 100 villages in total (23).
8. **ANM (Auxiliary Nurse Midwife):**

An auxiliary nurse midwife is defined as someone who assists in the provision of maternal and newborn health care, particularly during childbirth but also in the prenatal and postpartum periods.

9. **Anganwadi worker:**

Under the ICDS, scheme one trained person is allotted to a population of 1000, to bridge the gap between the person and organized health care, and to focus on the health and educational needs of children aged 0-6 years. This person is the anganwadi worker.

10. **Knowledge:** Degree to which the community health worker has the theoretical or practical understanding of the understanding of the function and task assigned to him/her (11).
11. **Competencies:** Degree to which the community health worker has the skills necessary to carry out the tasks assigned to him/her (11).
12. ***Village Health Sanitation and Nutrition Committee:*** Was introduced by National Rural Health Mission in 2005, to ensure community participation at all levels, which include participation as beneficiaries, in supporting health activities, in implementing, and even in monitoring and action-based planning for health programmes (24).
13. ***Primary Health Centre*** : There should be 1 PHC for every 30,000 (plain areas) & 20,000 in (Hilly and tribal areas) .
14. ***Community Health Centre*:** Is the secondary level of contact between the community and the health facility. There should be 1 CHC for every 1,20,000 (plain area) & 80,000 in ( Hilly and tribal area) as per the norms of Indian Public Health Standards.
